# Burden of Depressive and Anxiety Disorders in Nepal, 1990-2017: An Analysis of Global Burden of Disease Data

**DOI:** 10.1101/2021.06.13.21258841

**Authors:** Deepa Kumari Bhatta, Kreeti Budhathoki, Kiran Paudel, Shishir Paudel, Sujan Babu Marhatta, Avinaya Banskota, Yadav Prasad Joshi

## Abstract

**Objectives:** This study aims to describe the burden of depressive and anxiety disorders in Nepal in terms of incidence, prevalence, YLDs, and DALYs by age and sex.

**Design:** An ecological study on disease burden.

**Methods:** We used the publicly available Global Burden of Disease (GBD) data from 1990 to 2017. The age and sex-specific incidence, prevalence rate, Years Lived with Disability (YLDs), and Disability Adjusted Life Years (DALYs) were examined among per 100,000 populations.

**Results:** Females had a higher prevalence of depressive 4094.4 (95% uncertainty interval [UI]: 3761.9-4470.3 per 100,000) and anxiety 4496.8 (95% UI: 4171.9-4837.9 per 100,000) disorders. The prevalence of major depressive disorders was comparatively higher in females (2766.7) than males (1822.9). Females also had higher YLDs for both depressive and anxiety disorders. In 2017, higher DALYs of anxiety disorder were found in the females of 45-49 years (630.1). Childhood sexual abuse was found to be the main risk factor for depressive disorder, contributing to 32.5 DALYs in both sexes. Bullying, victimization had contributed to 26.7 DALYs of anxiety.

**Conclusions:** The high burden of disorders in females would support in identifying major mental health challenges in Nepal and develop plans and various preventive, curative, and rehabilitative strategies.

**Strength and limitations of this study:**

- This is one of the few studies that have provided the burden of depressive and anxiety disorders in developing nations such as Nepal.
- The data used in this study were based on the GBD studies which utilize exhaustive information sources and comprehensive investigations.
- Estimation of the burden of diseases was restricted to standard epidemiological measures and other financial and social burdens were not considered.
- Due to use of secondary data and information which were only available publicly this may not representative of the whole nation.

## INTRODUCTION

Depressive and anxiety disorders are the most frequent mental disorders and are ranked among the top conditions causing disability worldwide.^1^ Major depressive disorders (MDDs) are one of the most common and serious mental disorders linked to the diminished role, functioning and quality of life, medical morbidity, and mortality.^2^ Dysthymia as a persistent depressive disorder is a chronic mood illness and often is more disabling than episodic major depression. The consequences of dysthymia are increasingly recognized as serious; they include severe functional impairment, increased morbidity from physical disease, and increased risk of suicide.^3^ Anxiety disorders refer to a group of mental illnesses and their burden is increasing globally.^4,5^

The global burden of disease study (GBD) 2013 had ranked depressive and anxiety disorders in the second and ninth-highest causes of Years Lived with Disability (YLDs) respectively in both developed and developing countries.^6^ The burdens of these disorders have been increasing gradually over the last 27 years from fourth in 1990 to third in 2017.^7^ Globally, the number of incident cases of depression increased from 172 million in 1990 to 258 million in 2017, representing an increase of 49.9% cases.^8^ In South Asia, depressive disorders remain a higher burden in females and older adults. Bangladesh had the highest (4.4%) age-standardized prevalence in 2016 followed by Nepal (4.0%), India (3.9%), Bhutan (3.7%), and least (3.0%) in Pakistan.^9^

In Nepal, the burden of depression and anxiety disorders in terms of morbidity and disability are overwhelmingly high and have the second-highest prevalence rate (301 per 1,000) of psychiatric morbidity in comparison to other South Asian countries.^10,11^ Mental health problems including depressive and anxiety disorders have a higher prevalence in females and adults of 40-49 years’ age group in the country.^10^. Institute for Health Metrics and Evaluation (IHME) 2017 report has ranked depressive disorder as the fourth cause of YLDs.^12 13^ This study aims to describe the burden of depressive and anxiety disorders in Nepal in terms of incidence, prevalence, YLDs, and Disability-adjusted life years (DALYs) by age and sex.

## METHODS

### Research Design

This is an ecological study on disease burden based on the secondary data obtained from for Health Metrics and Evaluation (IHME).

### Data

The data used in this study were publicly available on the Global Health website of the IHME repository. The IHME is a research institute working in the area of global health statistics, disease burden, and impact evaluation at the University of Washington, USA. The GBD uses all accessible data sources, including national surveys, and other epidemiological studies to estimate the burden, quantify the disease-specific health loss, injuries, and risk factors.^14^ In the GBD study, depressive and anxiety disorders were included under mental disorders and depressive disorder is further categorized as MDD and dysthymia.^12^ The metrics, data sources, and statistical modeling for depressive and anxiety disorders were described comprehensively in GBD 2017.^7, 12, 13^

### Statistical analysis

In the estimation of each depressive and anxiety disorder, epidemiological data were generated using a Bayesian meta-regression tool DisMod-MR 2.1. The different surveys and scientific publications from 1990 to 2017 were considered as the data source for the study. The YLDs were calculated by multiplying the prevalence and disability weight. The DALYs were estimated from the estimates of YLLs and YLDs. The YLDs were mainly used in the estimation of DALYs in anxiety and depressive disorders.^7, 13^ The estimation for exposure and attributable disease burden of risk factors was made from the comparative risk assessment framework. The distribution of risk exposure was estimated based on research methods such as randomized controlled trials, prospective cohorts, or case control studies. Disease burden attributable to risk factors was estimated by comparing the present exposure distribution to the theoretical minimum risk exposure level. Population attributable fraction of uncertainty was calculated for each pair of disease risk. In the final estimates, exposure estimate, relative risk, theoretical minimum risk distribution, and uncertainty in the first outcome rates were propagated.^13^

We selected the risk factors to which the burden of depressive and anxiety disorders could be attributed in GBD 2017 including intimate partner violence, childhood sexual abuse, and bullying victimization. The age-standardized and sex-specific prevalence and DALYs in 2017 for depressive and anxiety disorders along with annualized percentage change in DALYs of all ages and both sexes for Nepal were estimated. We also compared the incidence and DALYs of depressive and anxiety disorders from 1990 to 2017 in age-standardized and sex-specific populations. Finally, the DALYs for depressive and anxiety disorders attributable to selected risk factors in 2017 were also estimated. The analysis of data was performed using Microsoft Excel.

### Patient and Public Involvement

There were no patients and public involvement in any aspect of this study as this study is based on secondary data.

## RESULTS

In 2017, the overall prevalence of the depressive disorder in females was 4094.4 (95% uncertainty interval [UI]: 3761.9-4470.3) per 100,000 and in males was 2847.2 (95% UI: 2614.6-3118.4) per 100,000. Likewise, the prevalence of anxiety disorder in females was 4496.8 (95% UI: 4171.9-4837.9) and in males was 2966.2 (95% UI: 2764.7-3178.6). Prevalence of both disorders was found comparatively higher in females than males. Females also had higher YLDs for depressive disorder. Likewise, The MDD prevalence was also found higher in females (females: 2766.7, males: 1822.9). For depressive disorders, a marked variation was observed in the YLDs in both sexes. The annualized rates of change in DALYs from 1990 to 2017 for both sexes and all ages were observed at 0.8% and 0.6% in depressive and anxiety disorders, respectively (table 1).

**Table 1.** Age-standardized prevalence and YLDs per 100,000 in 2017

| Diseases | Total Prevalence (95% UI) |  | Total YLDs (95% UI) |  | Annualized % change in DALYs |
| --- | --- | --- | --- | --- | --- |
|  | Male | Female | Male | Female |  |
| Depressive disorders | 2847.2 (2614.6-3118.4 ) | 4094.4 (3761.9-4470.3) | 468.5 (331.6-640.9) | 683.3 (483.0-923.1) | 0.8 |
| MDD | 1822.9 (1645.0-2028.5) | 2766.7 (2487.1-3081.6) | 365.3 (253.8-497.0) | 547.4 (384.4-739.8) | 0.8 |
| Dysthymia | 1074.5 (930.3-1246.1) | 1428.3 (1238.3-1647.4) | 103.1 (69.2-148.3) | 135.9 (91.2-198.5) | 0.8 |
| Anxiety disorders | 2966.2 (2764.7-3178.6) | 4496.8 (4171.9-4837.9) | 283.8 (201.7-378.5) | 425.5 (300.6-564.5) | 0.6 |

In between 1990 and 2017, the highest incidence and DALYs were found for depressive disorder in females. In males, the incidence of anxiety disorder remained stable after 2005 while the DALYs were found in increasing trend (figure 1).

**Figure 1:**
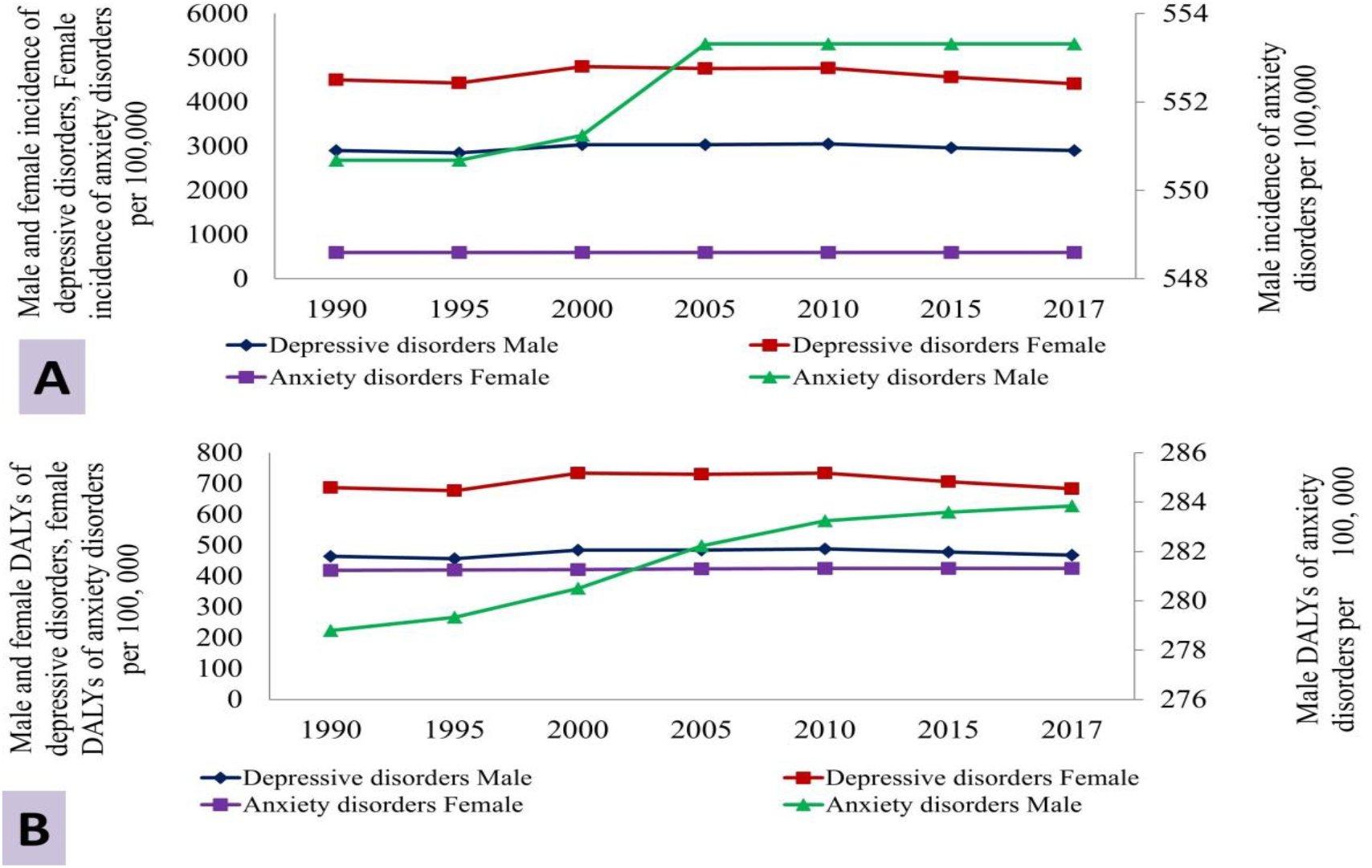
Age-standardized incidence (A) and DALYs (B) by sex from 1990-2017

Incidence and DALYs of depressive disorders illustrated the pattern of growth with age and peaks among the elderly population of 65 years and more and older age groups in both sexes. But surprisingly cases of anxiety disorder were found highest among 30-49 years and declined with increasing age in both sexes. Higher DALYs for anxiety disorders in both males (408.8 per 100,000) and females (630.1 per 100,000) were seen within the 45-49 years age group. In all age groups, females contributed more DALYs for both disorders than males (figure 2).

**Figure 2:**
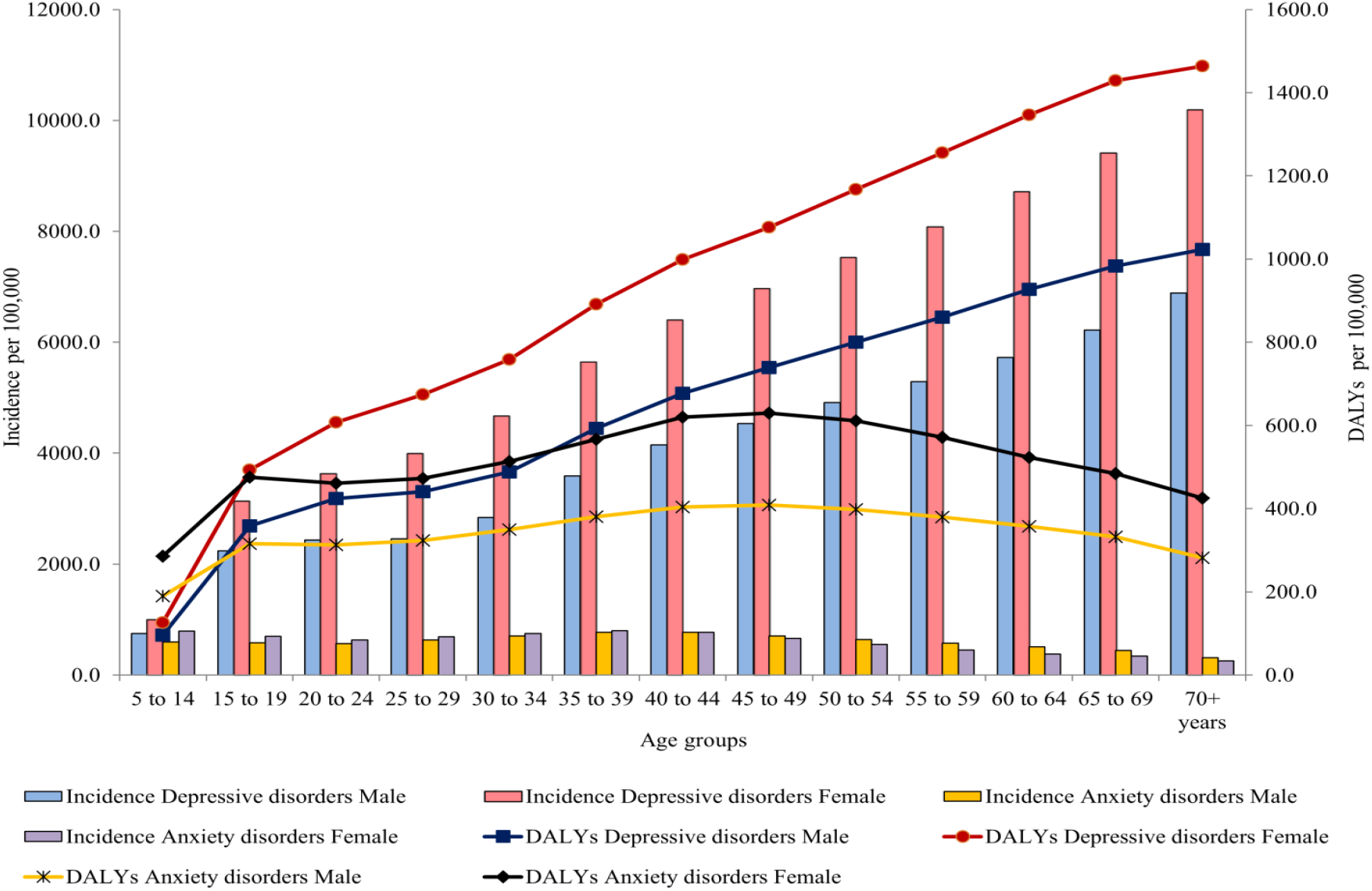
Age and sex-wise burden of depressive and anxiety disorder in 2017

In 2017, childhood sexual abuse was the main risk factor contributing to 32.5 DALYs for depressive disorder among both sexes followed by intimate partner violence (26.8) and bullying victimization (20.7) in 100,000 populations. Bullying victimization was the only risk quantified for anxiety disorders in GBD, representing 26.7 DALYs (figure 3).

**Figure 3.**
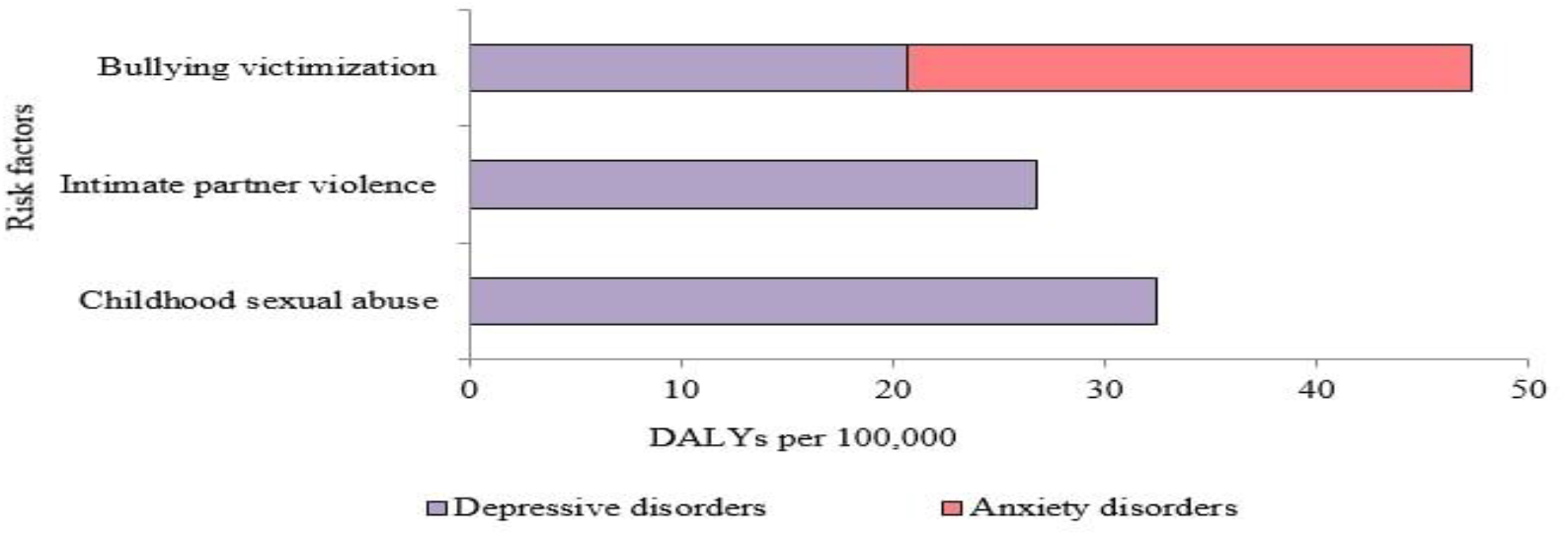
Age-standardized DALYs rate of depressive and anxiety disorders attributable to risk factors

## DISCUSSION

Globally, depressive and anxiety disorders are of utmost concern to public health contributing to global disabilities.^4^ In the 2004 estimates, MDD was considered to become the leading cause of global disability by 2030.^15^ However, the 2018 study reported that MDD has become the main cause of disability.^12^. These disabilities interfere with the individual’s economic and social productivity.^16^ The burden of both disorders is higher in developing and least developed countries compared to developed nations. In Nepal, depressive and anxiety disorders are of major health concern and contribute significantly to the loss of well-being, specifically in adults and older adults.^12^ The impacts of mental health disease burden are found in the development and prosperity of nations. Nepal, being one of the least developed countries has also realized depression and anxiety disorders as a serious public health issue.

Several previous studies conducted in the South Asia region including Nepal reported a higher prevalence of depression and anxiety disorders in females.^10,17-20^ The risks associated with higher mental disorders in females are linked to gender discrimination, widowhood, physical and sexual abuse, domestic and intimate partner violence, antenatal and postnatal stress, and adverse cultural norms including child marriages and the dowry system.^21^ Furthermore, armed conflict and political instability, low financial status, high vulnerability to natural disasters such as earthquakes, floods, and landslides have contributed to an increase in cases of mental disorder in the country.^17, 18, 22^ Similarly, studies from other high-income countries such as China and Singapore also found a higher prevalence of depressive and anxiety disorders in females.^23, 24^

Globally, the cases of depressive and anxiety disorders were increased by 49.9% in between 1990 to 2017.^8^ Our study showed an increase in incidence and DALYs of depressive disorders in Nepal until 2010 and a gradual decrease thereafter. The armed conflict and political instability of the country over 10-years remained one of the causes of mental distress in Nepal.^17,18, 22^ When disaggregated according to age, incidence and DALYs for depression increases in the elderly population. The finding is consistent with other studies in Nepal, India, and Pakistan.^19, 20, 22^ The possible associated factors might be distorted by traditional family structures, loneliness, and co-morbidities. Depressive and anxiety disorders were the predominant mental health problems with higher occurrence among adults of 40-49 years.^10^ Inadequate education, low income, personality traits, reduced social functioning, unhealthy lifestyle, and poor health are some reasons for mental health issues.^25^ In contrast, high income countries have not followed the pattern of our findings of depressive disorder burden increasing with age.^26, 27^. In young and adolescents, anxiety disorder has been increasing in Nepal and India.^20, 24,28^ The risk factors are low household income, gender discrimination, bullying, social media, family history, temperament, and personality.^25^

In various countries, childhood sexual abuse, intimate partner violence, and bullying victimization have increased the burden of depressive disorders in both sexes.^29, 30^ In our study, bullying victimization is the main risk factor for anxiety disorders. The finding is consistent with the GBD study of India.^20^ Besides, low self-esteem, family history of depression, female sex, number of traumatic experiences, and disturbing family environment are also associated risk factors for depressive and anxiety disorders.^28,31-33^

### Limitation of the Study

Although this study is one of the few studies that have provided the burden of depressive and anxiety disorders in developing nations using the data based on the GBD studies which utilize exhaustive information sources and comprehensive investigations, this study has its limitations. The secondary data and information might be based on data derived from publicly available sources that may not representative of the whole nation. Estimation of the burden of diseases was restricted to standard epidemiological measures and other financial and social burdens were not considered. Additionally, risk factors of depressive and anxiety disorders were not evaluated. Avoiding major estimators of the disease burden might risk genuine representation.

## CONCLUSIONS

The persistent burden of depressive and anxiety disorders from 1990 to 2017 remains one of the topmost causes of disabilities in the Nepalese population predominantly among females. The burden varies by age, peaking in older adults for depressive disorders and middle-aged for anxiety disorders. A comprehensive integrated social, environmental and behavioral approach along with investment in mental health services to provide affordable treatment, care, and rehabilitation is urgent and essential to address the escalating problem of depressive and anxiety disorders.

## Data Availability

The data used was of Global Burden of Disease (GBD)

## Contributions

DKB: Contributed in conceptualization, methodology, formal analysis, investigation, data curation. KB: Contributed in conceptualization, methodology, investigation. KP: Contributed in investigation, data curation. SP: Contributed in manuscript preparation, proofread and validation. SBM: Contributed in methodology, formal analysis AB: Contributed in investigation, proofread and validation. YPJ: Contributed in conceptualization, methodology, formal analysis, data curation. All authors participated in the manuscript review and approval.

## Data availability statement

The data required to calculate disease burden were publicly available in the homepage of Institute of Health Matrices and Evaluation (IHME), USA. The data are available upon request.

## Ethical approval

Not required

